# Investigations on druggable gene mutations related to AML/ALL lineage genes in Advanced Phases of CML: Implications in patient-tailored therapy of blast crisis CML in TKI era

**DOI:** 10.1101/2024.09.08.24313260

**Authors:** Nawaf Alanazi, Abdulkareem AlGarni, Sarah AlMukhaylid, Maryam AlMajed, Sabreen Alanazi, Muhammad Aamir Khan, Muhammad Farooq Sabar, Mudassar Iqbal, Abid Jameel, Akhtar Hussian, Dhay Almaghlouth, Alhanoof Alsuwaidani, Ghala Basem Alsalem, Nouf AlMutairi, Hassan H Almasoudi, Sarah Alfaye, Maryam Butwyibah, Batool Salman Alnajad, Fatimah Hussain Alali, Anwar Hussain Al-Rasasi, Fatimah Ali Alabdullah, Kanza Adeel, Sara Faisal Alfayez, Tarig Karar, Fahad M. Alsaab, Yaqob Samir Taleb, Noran Aboalela, Sana Shahbaz, Sumyiea Riaz Malik, Amer Mahmood, Sulman Basit, Muhammad Anharullah, Buthainah AlShehab, Sultan Al-Qahtani, Abdullah Alruwaili, Mahmood Rasool, Muhammad Asif, Aamer Aleem, Irtaza Fatima Zafar, Rizwan Naeem, Masood A. Shamas, Zafar Iqbal

**Author notes:** Authors contributed equally and share FIRST AUTHORSHIP (Nawaf Alanazi, Abdulkareem AlGarni, Sarah AlMukhaylid, Maryam AlMajed, Sabreen Alanazi, Dhay Almaghlouth, Sara Faisal Alfayez, Zafar Iqbal). (www.pak-hematology.org). Correspondence: Prof. Dr. Zafar Iqbal.

## Abstract

**Background:** Chronic Myeloid Leukemia (CML) is a myeloproliferative stem cell malignancy. Chronic Phase CML (CP-CML) is treatable with overall survival equivalent to the general population. Nevertheless, a proportion of CP-CML progresses to the accelerated phase (AP-CML) and ultimately blast crisis (BC-CML), with the latter having an overall survival (OS) of 3-23 months, making it almost a fatal manifestation. Therefore, the treatment of BC-CML is one of the biggest challenges in modern cancer medicine. Moreover, the OS of BC-CML is very variable indicating its heterogeneity. Although BC-CML is a different clinical entity than acute leukemias, it resembles AML (as myeloid BC-CML) or ALL (lymphoid BC-CML). Therefore, this study was designed to find out AML-/ALL lineage gene mutations in BC-CML using very sensitive next-generation sequencing.

**Patients & Methods:** The study included 141 CML patients (123 CP-CML as control groups; 6 AP-CML and 12 BC-CML as experimental groups). Most of the patients received imatinib mesylate (IM) as first-line treatment. All response criteria were per European LeukemiaNet (ELN) guidelines 2020. Whole exome sequencing (WES) was carried out to find out druggable gene mutations and the druggability of the mutated genes was determined using the online Artificial intelligence (AI) tool www.pandrugs.com. SAS/STAT software version 9.4 was used for data analysis (SAS Institute Inc., Cary, NC, USA). For statistical computing, the R package was employed (Vienna, Austria). The study was approved by the ethical committee of KAIMRC and carried out per the guidelines of the Helsinki Declaration

**Results:** Overall male-to-female ratio was 1.6:1 and the mean age was 36.4 (range: 9-67) years. Eighteen (12.8%) patients progressed to AP-CML while 12 (8.5%) to BC-CML finally. BC-ML patients had overall poorer response to TKIs and higher mortality rate (75%) that prompted to look for additional gene mutations. WES showed overall 64 AML**-**/ALL-associated gene mutated in advanced phase CML patients. Overall WES coverage was about 110X. AP-CML had 1644 variants, whereas BC-CML had 2531 variants, with a 54% gain in mutations from AP-CML to BC-CML (P< 0.000001). Among AML-/ALL-related mutated genes were NPM1 (%1.98), DNMT3A (%1.86), PML (%1.82), AKT1 (%1.62), CBL (%1.30), JAK2 (%0.71), TET2 (%0.59), IDH1 (%0.32), and BCL2.

**Conclusions:** NGS analysis of AP-& BC-CML found mutations in many AML-/ALL-lineage genes, which is much higher than previously reported. This shows a huge genetic similarity between BC-CML and AML/ALL. FDA-approved and various novel experimental drugs under clinical trials are available for some of these genes we reported in this study. We conclude that our approach can help in finding druggable gene mutations related to AML-/ALL-lineage genes in almost every BC-CML patients and provide a practical guidance for drug repurposing as well as gateway to in-trail novel experimental drugs to individualize BC-CML patient treatment.

**Tweetable Abstract:** Blast crisis Chronic Myeloid Leukemia (BC-CML) is fatal due to its limited number of therapeutic options due to its clinical and genetic heterogeneity. In this study, we report AML-/ALL-lineage gene mutations associated with BC-CML, their implications in further comprehending BC-CML biology in clinical management.

## Introduction

Chronic Myeloid Leukemia (CML) is a myeloproliferative disorder that arises from the abnormal proliferation of cloned hematopoietic stem cells (HSC), leading to the excessive production of non-functional cells, particularly granulocytes, in both the bone marrow and peripheral blood (1). Philadelphia chromosome t(9;22), which is a hallmark of CML, arises from a reciprocal translocation involving the BCR on chromosome 22 and ABL on chromosome 9. This fusion leads to the constitutive expression of the tyrosine kinase BCR-ABL on HSCs, causing the continuous proliferation and growth of leukemic stem cells (LSCs) (2). The global incidence of CML is 1-2 cases per 100,000 adults, constituting approximately 15% of new adult leukemia cases (3).

CML undergoes a triphasic progression, initiating with the indolent Chronic Phase (CP) that is marked by a notable increase in myeloid precursors and mature cells, with overall survival equal to the general population, at least in technologically advanced countries, due to the introduction of tyrosine kinase inhibitors (TKIs) in last three decades (4). However, a subset of CML patients progress to the accelerated phase (AP) and ultimately blast crisis (BC), the later having an overall survival of 3-18 months (3). The Blast Crisis is distinguished by a rapid proliferation of primitive cells in both the bone marrow and blood, treatment failure, drug resistance, relapses and ultimate death (5). It makes BC-CML treatment one of the biggest challenges in modern cancer medicine in the 21^st^ century. Therefore, there is a dire need to find new drug targets and therapies for BC-CML (1, 5).

Typically, BC-CML resembles either Acute Myeloid Leukemia (AML) or Acute Lymphoid Leukemia (ALL), referred to as myeloid blast crisis (M-BC) or lymphoid blast crisis (L-BC) (6). A large number of genes related to myeloid and lymphoid lineage have been reported to be mutated in AML and ALL patients and FDA-approved inhibitors against these gene mutations are effectively being utilized in routine clinical practice (7, 8). However, it remains to be elucidated if the same AML and ALL associated genes may be responsible for myeloid and lymphoid blast crisis, respectively, that can provide the opportunity to treat MBC-CML and LBC-CML patients using FDA-approved drugs developed against AML and ALL lineage genes. As no studies have been carried out previously in this regard, the objective of this study was to find out AML and ALL lineage gene mutations in blast crisis CML patients and create a database of FDA-approved and experimental drugs developed against AML and ALL specific gene mutations that have the potential to be utilized for treating BC-CML.

## Methodology

### Patient Inclusion and Exclusion Criteria

This research comprised 141 CML patients from Hayatabad Medical Complex (HMC), Peshawar, Pakistan from January 2012 to December 2023. All patients were given imatinib mesylate (IM) as first-line treatment and those who developed IM resistance were given nilotinib (NI). Out of 141, 123 age/gender-matched Chronic Phase (CP-CML) patients served as controls, while 12 AP-CML and 6 BC-CML individuals with Accelerated Phase (AP-CML) were included in the experimental group. The European Leukemia Net (ELN) recommendations from 2013 and later on ELN 2020 were used for all clinical classification and treatment response criteria (9–11).

### Definitions of Clinical Phases of Chronic Myeloid Leukemia (CML) for Staging

#### Chronic phase (CP)

A CML case was established as chronic phase (CP) if it was detected in the blood that blast cells were less than 5%, basophils were 15-19%, blasts and promyelocytes were less than 30% and there were no blast cells in extramedullary areas (9).

#### Accelerated phase (AP)

The criteria for establishing an Accelerated phase (AP) was as follows: the presence of blasts was approximately 15-29%, the presence of promyelocytes and blasts accounted for more than 30% or more in the blood or the bone marrow, a low platelet count (100×10^9^/L), a basophil presence of 20% or greater and Ph+ cells with chromosomal anomalies (9).

#### Blast Crisis (BC)

The criteria for classifying a Blast Crisis phase (BC) was as follows: the presence of 30% or more of blasts in either the blood or the bone marrow along with in extramedullary sites (9).

### Criteria for Assessment of Treatment Response in Chronic Myeloid Leukemia

To monitor the therapy response, blood counts and physicals were monitored on a monthly basis. The three response tools listed below were used to assess the drug’s efficacy in CML patients (9).

#### Complete Hematological Response (CHR)

It was identified by a complete lack of immature cells and palpation in the spleen along with the drop to normal counts of WBCs (< 10×10^9^/L), basophils (< 5%) and platelets (< 450×10^9^/L) (9).

#### Cytogenetic Response (CyR)

Testing bone marrow aspiration at diagnosis 6 months, 12 months and yearly in order to evaluate the cytogenetic response and the differential morphology. A complete cytogenetic response (CCyR) is indicated if FISH detected <1% BCR-ABL nuclei (Ph+) per 200 cells. A partial cytogenetic response (PCyR) was classified by the occurrence of Ph+ in approximately 1-35%, while minor cytogenetic response (mCyR) was 36-65% Ph+. If Ph+ was determined as 66-95% then it was categorized as the minimal cytogenetic response (miCyR), whereas the no cytogenetic response (nCyR) was categorized when Ph+ was detected as 95% or above (9).

#### Molecular Response (MR)

Major molecular response (MMR) was defined if the BCR-ABL/ABL ratio threshold was 0.1% or lower. MR^4^ and MR^4.5^ had a ratio of ≤ 0.01% and ≤ 0.0032%, accordingly (9). It was updated according to ELN 2020 later on (11).

#### Deep Molecular Response

Deep Molecular Response (DMR) is termed as the detection of *BCR::ABL1* transcripts at less than 0.01% using RT-qPCR, making such patients in Treatment Free Remission (TFR), around half of which detected later a *BCR::ABL1* transcripts level that is higher than 0.1% (12).

### Criteria for Calculation of European Leukemia Net (ELN) Responses and Survival

#### Optimal response

This was determined if the following was observed: a decrease in the Ph+ to ≤35% at three months then by six months it was absent. After 12 months and at any point afterward, BCR-ABL1 PCR should be 0.1% (9).

#### Warning

It was thought to be a warning if at diagnosis there was a substantial risk along with CCA/Ph+ major route, followed by Ph+ levels to 36-95% at three months and 1-35% at six months and twelve months if PCR of BCR-ABL1 was greater than 0.1-1% (9).

#### Failure

It was deemed as a failure if there was a lack of CHR or CCyR and if at three months Ph+ was greater than 95%, at six months it was more than 35% and at twelve months it was more than 0% (9).

## Criteria for Documenting Adverse Events

Adverse events were categorized by standard terminology version 4.03 (13).

## Ethical Approval

The protocol for the study was approved by King Abdullah International Medical Research Center (KAIMRC) and King Saud bin Abdulaziz University for Health Sciences (KSAU-HS) Saudi Arabia, Hayatabad Medical Complex (HMC) and University of the Punjab, Lahore, Pakistan. All patients who took part in the trial provided signed informed consent. Throughout this study, the requirements of the Helsinki Declaration were implemented (**General Assembly of the World Medical Association).**

## Sample Collection and DNA Extraction

Peripheral blood samples were collected from study subjects using EDTA tubes with a volume of around 3-5ml. These samples were obtained during the biweekly visits to the outpatient department of the medical oncology unit at HMC. The samples were stored at −70°C for further examination. Before DNA extraction, the patient samples, as well as the DNA extraction and reagents were warmed to room temperature (15-25°C).

The DNA extraction consists of several steps. It involved the mixing of 200 microliters of the blood sample, 200 microliters of Buffer AL and 20 microliters of QIAGEN Protease in a tube. The resultant mixture was incubated for 10 minutes at 56°C after it was vortexed. 200 microliters of ethanol was added and the mixture was vortexed again, then it was transferred to a QIAamp Mini spin column and the extract was removed after centrifugation. After that, the flow-through was removed after the addition of Buffer AW1 and the centrifugation, repeating the procedure with Buffer AW2 to ensure the removal of all buffers, the column was centrifuged. The DNA was eluted using distilled water or adding Buffer AE and it was later incubated and centrifuged. At 260 nanometers, absorbance was measured to detect the concentration of DNA. If the concentration had a ratio of 1.7-1.9 at A260/A280, then it was considered pure. Using a Nanodrop Spectrophotometer the DNA was quantitated. The DNA was diluted into 70-80 ng/µl and 40 ng/µl to be used for WES and Sanger Sequencing respectively. Until further test, the DNA was stored at −80°C.

## Targeted Next Generation Sequencing (NGS)

CML samples, corresponding with the advanced clinical phases of the CML and an equal number of CP-CML as controls were processed for Next Generation Sequencing (NGS). To capture the extracted DNA, exome-capturing arrays were used following the DNA extraction (14). The SureSelectXT V6-Post Capture Exome kit was used to construct the library and enrich the exomes following DNA capture. The SureSelect kit encompassed about 99% of the reference sequences database and 214,405 exons containing splice sites (15). Following DNA fragmentation, tagmentation of the DNA fragments was done along with the amplification and purification, which were done by magnetic beads. To capture the designated regions, oligos were used. PCR was used to enlarge the enriched libraries, which were then measured using a Qubit fluorometer. The Agilent Bioanalyzer was set up to determine the library size dispersion. The Illumina NextSeq500 equipment was used to conduct cluster generation and full exome sequencing by depositing quantifiable DNA libraries into the flow cell (15).

## Next Generation Sequencing (NGS) Data Analysis

To transform the BCL files into FASTQ files, the BCL2FASTQ program was used. BWA Aligner was employed to align the FASTQ data to the human genome by the BWA-MEM algorithm. The annotation and filtration of genomic variants, which were called by the Genome analysis tool kit (GATK) was done using Illumina Variant Studio (15).

## Primary Analysis

To identify a shared biomarker for CML growth, mutated genes were analyzed in all advanced-phase CML patients. Filtration strategies that relied on calling rare variants and excluding intron and synonymous variants were applied to modify the Excel file presenting NGS. Furthermore, all variants with known prediction were removed, either benign (B) or tolerant (T). Some variants were considered B when it had 70% or more of B, while others were classified as T when T’s frequency was 70% or more (16). Variants with more than 0.005 population frequency in the dbSNP and Exome Sequencing Project (ESP) database were also eliminated. Thus, variant calling was only limited to variants with intermediate and high protein effects along with splice variants, resulting in about 124 rare variants. Moreover, data was further analyzed to investigate novel gene mutations that are present in AP-CML patients but not in CP-CML patients and healthy controls, suggesting its role in disease progression (17, 18). Access to data made by next-generation sequencing can be obtained from NCBI, to which it was submitted at https://www.ncbi.nlm.nih.gov/sra/PRJNA734750 (SRA accession number PRJNA734750; accessed on 28 September 2022).

## Finding out druggable mutations of myeloid and lymphoid lineage (AML and ALL related) genes

Using the exome sequencing data, we sorted for the genes already reported mutated in AML and ALL, thus covering myeloid and lymphoid lineage genes. For this purpose, we created a list of genes mutated in AML and ALL using the literature from PubMed. Out of the genes in this list, genes exclusively mutated in AP-CML and BC-CML but not in CP-CML or healthy controls (taken from genomic databases) were sorted and processed for further analysis.

## Investigating druggability of AML/ALL lineage genes

It is significant to note that not all genes mutated in ALL/AML have druggable mutations. Therefore, online artificial intelligence (AI) tools were used to investigate the druggable gene mutations out of a sorted list of genes exclusively mutated in AP-CML and BC-CML but not in CP-CML or healthy controls. For this purpose, we used tools available at www.pandrugs.com to find out druggable gene mutations related to ALL/AML lineage in our group of AP-CML and BC-CML. The standard procedures were utilized for this purpose (19). Druggability was determined by searching for the gene names in pandrug2 database. The gene is considered druggable if any drug for this gene is FDA approved to treat AML or ALL.

## Statistical Analysis of Patient Clinical Data

Based on the normality test, absolute numbers and percentages were demonstrated for categorical variables; mean and an appropriate measure of variation were demonstrated for continuous variables. For categorical data, chi-square or Fisher’s exact test was used to compare two groups, while a two-sample independent test or Mann–Whitney U test was used for the continuous data. ANOVA or Kruskal–Wallis test was used to analyze variance for groups of ≥3. To assess the survival outcome, Kaplan-Meier survival analysis curves were plotted (20). The group comparison was performed by a log-rank test. SAS/STAT software version 9.4 was used for data analysis (SAS Institute Inc., Cary, NC, USA). For statistical computing, the R package was employed (Available from: https://www.pandrugs.org; Vienna, Austria). The Eutos risk score, Euro risk score and Sokal risk score were measured (16, 17, 21).

## Results

This study included 141 patients diagnosed with CML. Of the total cases, 4.3% (N=6) were AP-CML and 8.5% (N=12) were BC-CML. The study subjects comprised 62.4% (N=88) males and 39% (N=55) females, with a male-to-female ratio of 1.6:1. The mean age of all patients was 36.4, ranging from 9 to 67 years. Laboratory results demonstrated that 87.7% (N=111) of the patients developed anemia whereas 56% (N=79) developed leukocytosis with more than 50×10^9^/L WBC count. Splenomegaly was seen in 71.6% (N=101) of patients. Furthermore, a comparison between different phases of CML patients showed that leukocytosis was significantly increased in advanced phase CML as compared to CP-CML cases (P=0.0184) (Table 1). Regarding clinical assessment, a significant relationship was seen between hepatomegaly and advanced-phase CML patients (P=0.0014). Although splenomegaly was observed in all phases, it was significantly associated with advanced phase CML as all AP-CML and BC-CML patients developed splenomegaly compared to only 67.5% (N=83) of CP-CML patients (p=0.0130). Furthermore, the size of the spleen was significantly increasing with the disease progression (P=0.0134), Table 1.

**Table 1:** Comparison between demographics and clinical characteristics in different phases of CML.

| Characteristics | Patients Group |  |  | P-VALUE |
| --- | --- | --- | --- | --- |
|  | CP-CML, n (%) | AP-CML, n (%) | BC-CML, n (%) |  |
| No. of patients | 123 (87.2) | 6 (4.3) | 12 (8.5) |  |
| Age, years |  |  |  |  |
| Mean (range) | 35.5 (9-67) | 35.6 (27-43) | 38.1 (29-50) |  |
| Gender |  |  |  |  |
| Male | 74 (60.2) | 4 (66.67) | 8 (66.7) | P = 0.6004 |
| Female | 49 (39.8) | 2 (33.33) | 4 (33.3) | P = 0.5987 |
| P-VALUE | P = 0.0272 | P = 0.3980 | P = 0.2933 |  |
| Ratio: Male: Female | 1.5-1 | 2: 1 | 2: 1 |  |
| Hemoglobin (g/dL) Mean | 10.1 |  |  |  |
| <12g/dl | 69 (56.1) | 5 (83.3) | 9 (75) | P = 0.0642 |
| >12g/dl | 14 (11.4) | 1 (16.7) | 3 (25) | P = 0.2609 |
| P-VALUE | P = 0.0024 | P = 0.2154 | P = 0.1380 |  |
| WBC count ($\times 10^9/L$) Mean | 313.7 | 315 | 325 | |
| <50 | 20 (16.3) | 1 (20) | 2 (16.7) | P = 0.8276 |
| $\geq 50$ | 64 (52) | 5 (80) | 10 (83.3) | P = 0.0184 |
| P-VALUE | P = 0.0052 | P = 0.2752 | P = 0.0661 |  |
| Platelets ($\times 10^9/L$) Mean | 400.2 | | | |
| <450 | 75 (61) | 4 (66.7) | 10 (83.3) | P = 0.2528 |
| $\geq 450$ | 33 (26.8) | 2 (33.3) | 2 (16.7) | P = 0.8722 |
| P-VALUE | P = 0.0011 | P = 0.4786 | P = 0.0661 |  |
| Imatinib |  |  |  |  |
| Yes | 82 (66.7) | 4 (66.7) | 7 (58.3) | P = 0.7260 |
| Nilotinib as 2nd Line |  |  |  |  |
| Yes | 41 (33.3) | 4 (66.7) | 8 (66.7) | P = 0.0065 |
| Hydroxyurea |  |  |  |  |
| Yes | 82 (66.7) | 3 (50) | 10 (83.3) | P = 0.9967 |
| Interferon |  |  |  |  |
| Yes | 41 (33.3) | 0 (0) | 0 (0) | P = 0.0038 |
| Chemotherapy |  |  |  |  |
| Yes | 0 | 0 | 9 (75%) | **P < 0.00001** |
| Splenomegaly |  |  |  |  |
| No | 40 (32.5) | 0 (0) | 0 (0) | **P = 0.0044** |
| Yes | 83 (67.5) | 6 (100) | 12 (100) | **P = 0.0130** |
| <5cm | 4 (3.3) | 0 (0) | 0 (0) | **P = 0.4358** |
| 5-8cm | 9 (7.3) | 1 (16.7) | 3 (25) | **P = 0.0619** |
| >8cm | 70 (56.9) | 5 (83.3) | 9 (75) | **P = 0.0732** |
| **P-VALUE** | **P = 0.0134** |  |  |  |
| Hepatomegaly |  |  |  |  |
| Yes | 35 (28.5) | 4 (66.7) | 8 (66.7) | **P = 0.0014** |
| Anemia |  |  |  |  |
| Yes | 97 (78.9) | 5 (83.3) | 9 (75) | **P = 0.9807** |
| Pregnant |  |  |  |  |
| Yes | 4 (8.2) | 0 (0) | 0 (0) | **P = 0.2090** |
| Survival Status |  |  |  |  |
| Confirmed deaths | 10 (8.1) | 0 (0) | 9 (75) | **P = 0.0003** |
| Alive at last follow up | 113 (91.9) | 6 (100) | 3(25) | **P = 0.0003** |

## Treatment outcomes

After using imatinib mesylate (IM) as the first-line treatment for all patients, 16.3% (N=20) of patients were either intolerant or did not respond to the drug even after increasing the dose. These 20 patients were treated with nilotinib (NI) as second-line therapy, 60% (N=12) of which were CP-CML, 10% (N=2) were AP-CML and 30% (N=6) were BC-CML patients. Out of the BC-CML patients receiving NI (N=6), 66.7% (N=4) were non-responders to the second-line treatment. All BC-CML patients were treated as Philadelphia-positive acute leukemia patients. There was a significantly high mortality rate of 75% (9/12) for BC-CML patients compared to CP-CML patients, who had a comparatively very low mortality rate of 8.1% (10/123), with one death not related to cancer. In addition, the overall survival rates for CP-CML and AP-CML patients were 91.9% (113/123) and 100% (6/6), respectively, whereas BC-CML patients had a survival rate of 25% (3/12). Our data shows the limited effectiveness of current treatment modalities to treat BC-CML and thus necessitates a hunt for new drug targets and novel treatments. It also highlighted the necessity to find early biomarkers for CML progression that could be targeted to prevent progression.

## Whole Exome Sequencing (WES)

As no specific biomarkers exist for CML disease progression and early detection of the patient group at risk of disease progression and most of the BC-CML patients specifically showed resistance to all drugs indicating non-availability of the effective drugs for BC-CML, advanced phase CML patient samples were subjected to whole exome sequencing (WES) to find out druggable mutations. The sequenced samples constituted 33.3% (N=5) AP-CML, 46.7% (N=7) BC-CML and 20% CP-CML samples. WES detected numerous variants in the above-mentioned study subjects. We included genes that were only mutated in advanced phases CML patients, i.e. AP-CML and BC-CML, but not in CP-CML patients and healthy control DNA sequences taken from genomic databases.

The total number of variants found in our advanced phases sequenced samples was 4175 (Figure 1). The mutated gene with the highest variant frequency in both AP-CML and BC-CML patients was RPTOR (7.3%), followed by BRCA1/BRCA2 (7.0%), BCR (6.0%), STAB1 (4.6%), NF1 (4.4%), ACIN1 (4.4%), EGFR (3.9%), NDRG2 (3.7%), ERG (3.3%) and MYH11 (3.1%). The number of variants and their frequencies in each gene were compared between AP-CML and BC-CML (Figure 2). AP-CML samples had 1644 variants, whereas BC-CML samples had 2531 variants, with a 54% gain in mutations from AP-CML to BC-CML (P< 0.000001) (Figure 2). The gain of mutation from AP-CML to BC-CML in the genes with the highest variant frequency was also significantly high, with the highest increase percentage of 79.3% observed in EGFR, followed by BRCA1/BRCA2 (72.9%), ACIN1 (69.1%), NF1 (60.6%), STAB1 (59.5%), MYH11 (58.0%), RPTOR (56.3%), ERG (46.4%), NDRG2(36.9%) and BCR (26.4%). Moreover, the low-frequency genes had a significant (52.5%) gain in variants from 866 variants in AP-CML to 1321 variants in BC-CML (P< 0.00001) (Figure 2).

**Figure 1:**
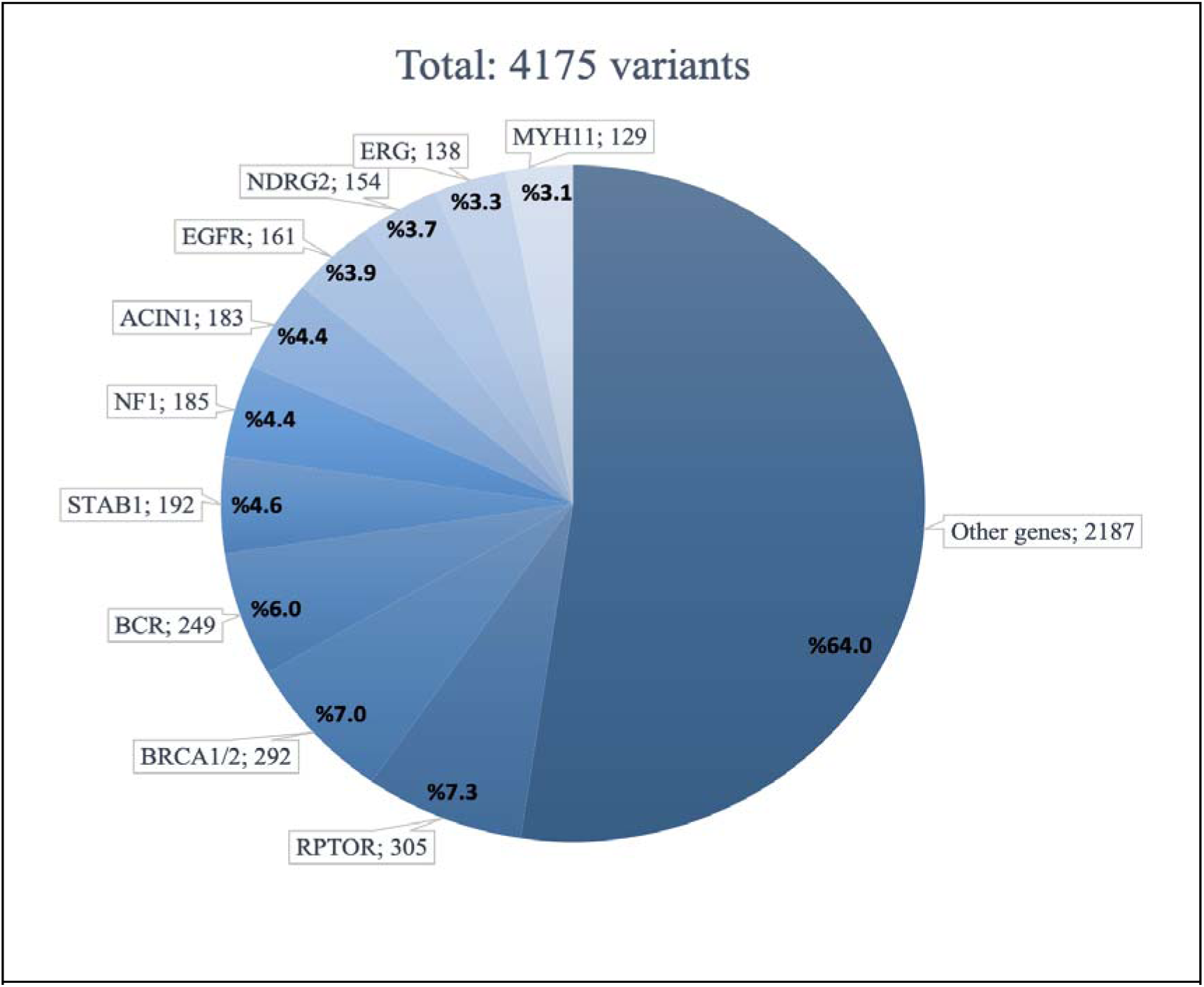
Variant frequencies AP-/BC-CML patient samples.

**Figure 2:**
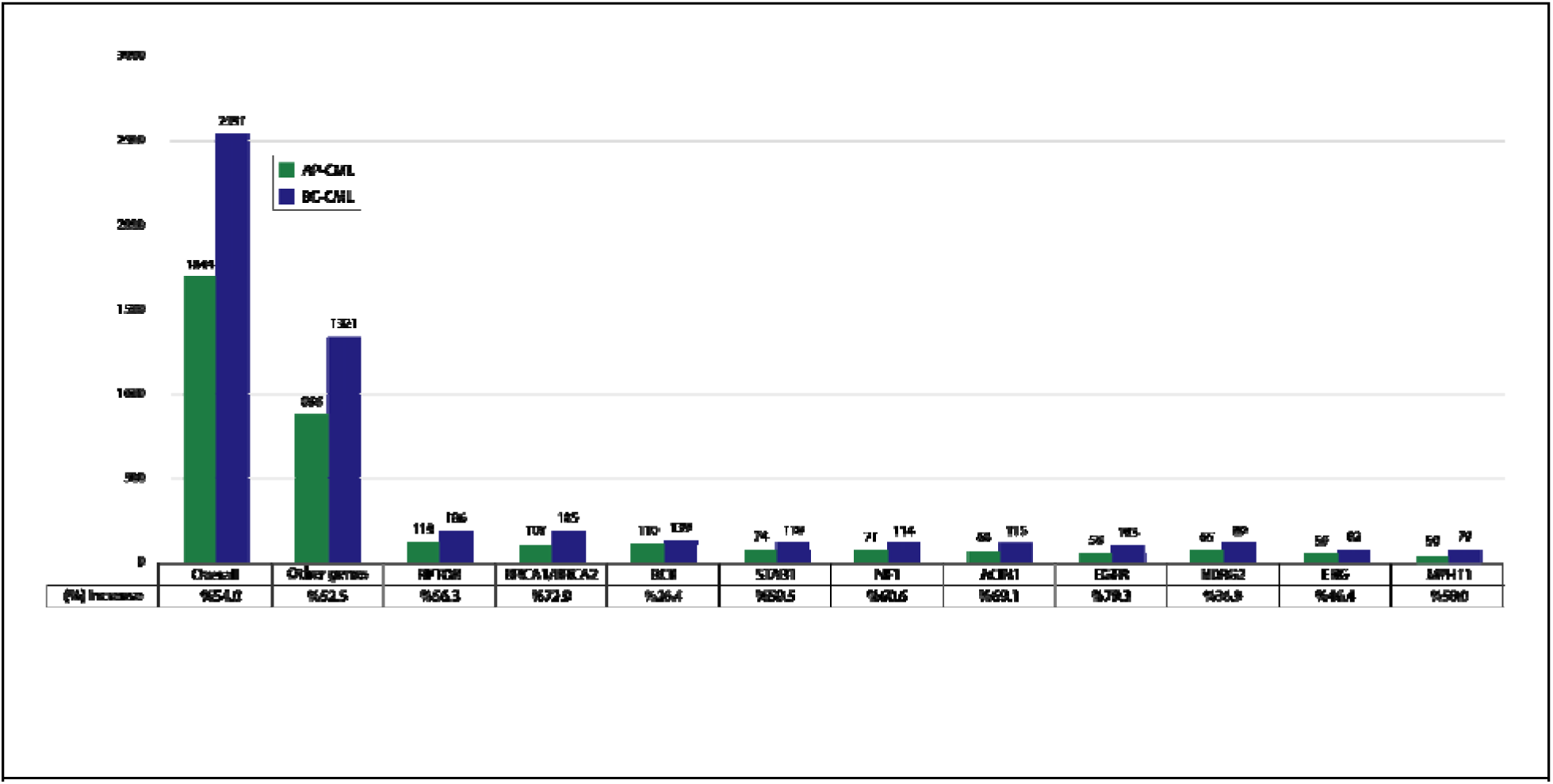
A comparison of the variants’ percentage increase from AP-CML to BC-CML in the highest variant frequency genes found in our study subjects.

## Druggability of mutations and drug repurposing

In the current study, we focused on druggable genes, regardless of their variant frequency. A gene is considered druggable if any drug for this gene was indicated to treat AML in the pandrugs2 database and literature, either as target therapy or chemotherapy. From the 64 mutated genes detected in our BC-CML patients, there were 9 druggable genes that are FDA approved for either AML or ALL patients. These druggable genes had low variant frequencies among our BC-CML patient samples.

The druggable gene with the highest variant frequency was NPM1 (1.98%), followed by DNMT3A (1.86%), PML (1.82%), AKT1 (1.62%), CBL (1.30%), JAK2 (0.71%), TET2 (0.59%), IDH1 (0.32%) and BCL2 (0.20%) (Figure 3). Although these genes had low variant frequencies in our study subjects, their variants showed a high percentage increase from AP-CML to BC-CML, indicating their significant role in disease progression (Figure 4). Almost all of these genes had targeted drugs that were FDA approved for the treatment of AML patients in general, except for IDH1, which had an FDA approved drug for IDH1 positive AML patients (Table 2). The FDA approved drugs are Arsenic Trioxide, Venetoclax, Doxorubicin, Mitoxantrone, Tretinoin, Quizartinib, Decitabine, Azacitidine and Ivosidenib. On the other hand, BCL2 was the only gene that had FDA approved drugs to treat ALL patients, Doxorubicin and Vincristine (Table 2). In addition, drugs approved for non-targeted AML or ALL treatment showed effectiveness as targeted therapy in clinical trials and experimental studies (Table 2). Drugs that are FDA approved for other types of leukemia are categorized according to their type of interaction with the genes, direct target (Table 3) and biomarker (Table 4).

**Figure 3:**
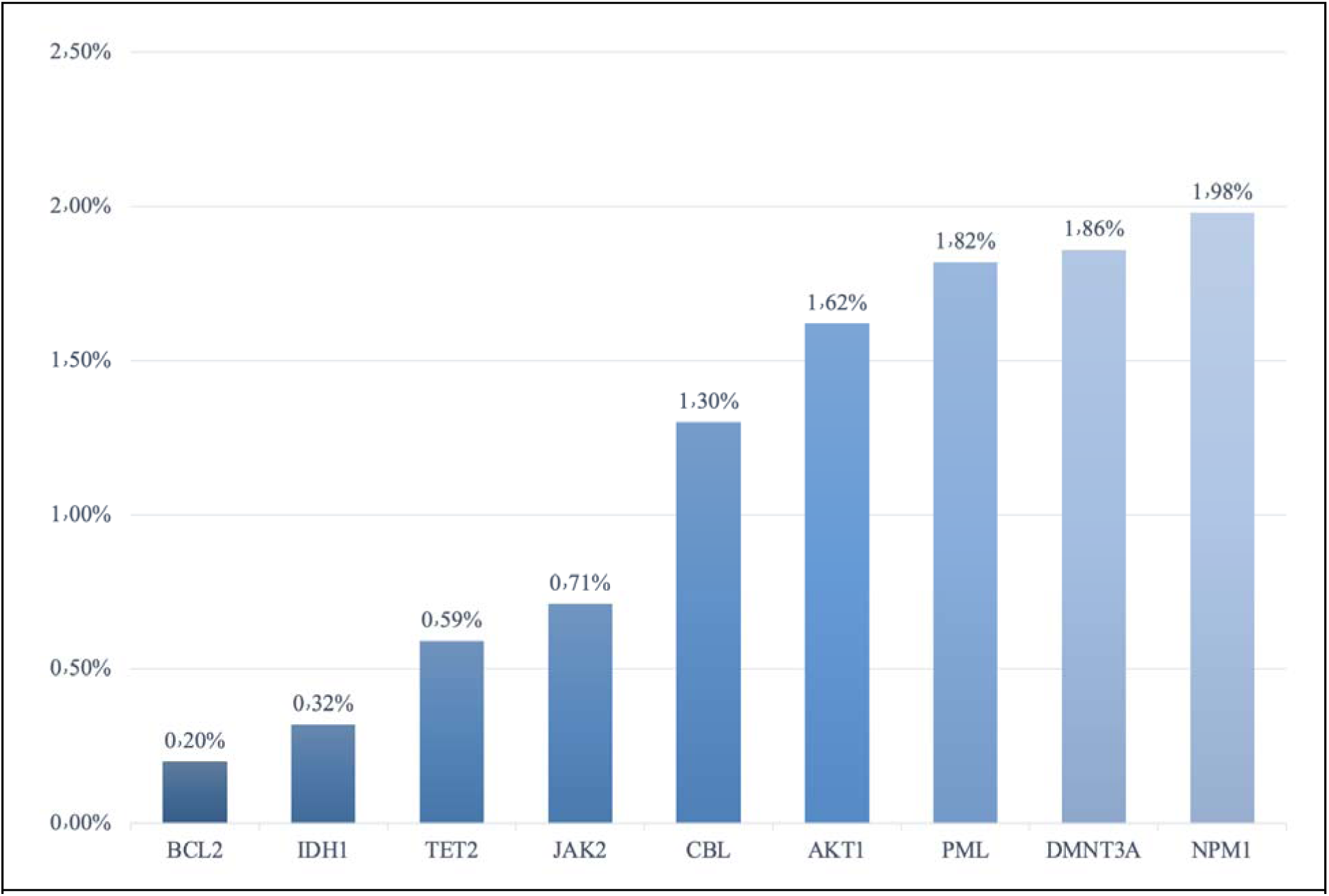
Variant frequencies of the druggable genes found in our BC-CML study subjects, which have FDA approved drugs to treat AML or ALL.

**Figure 4:**
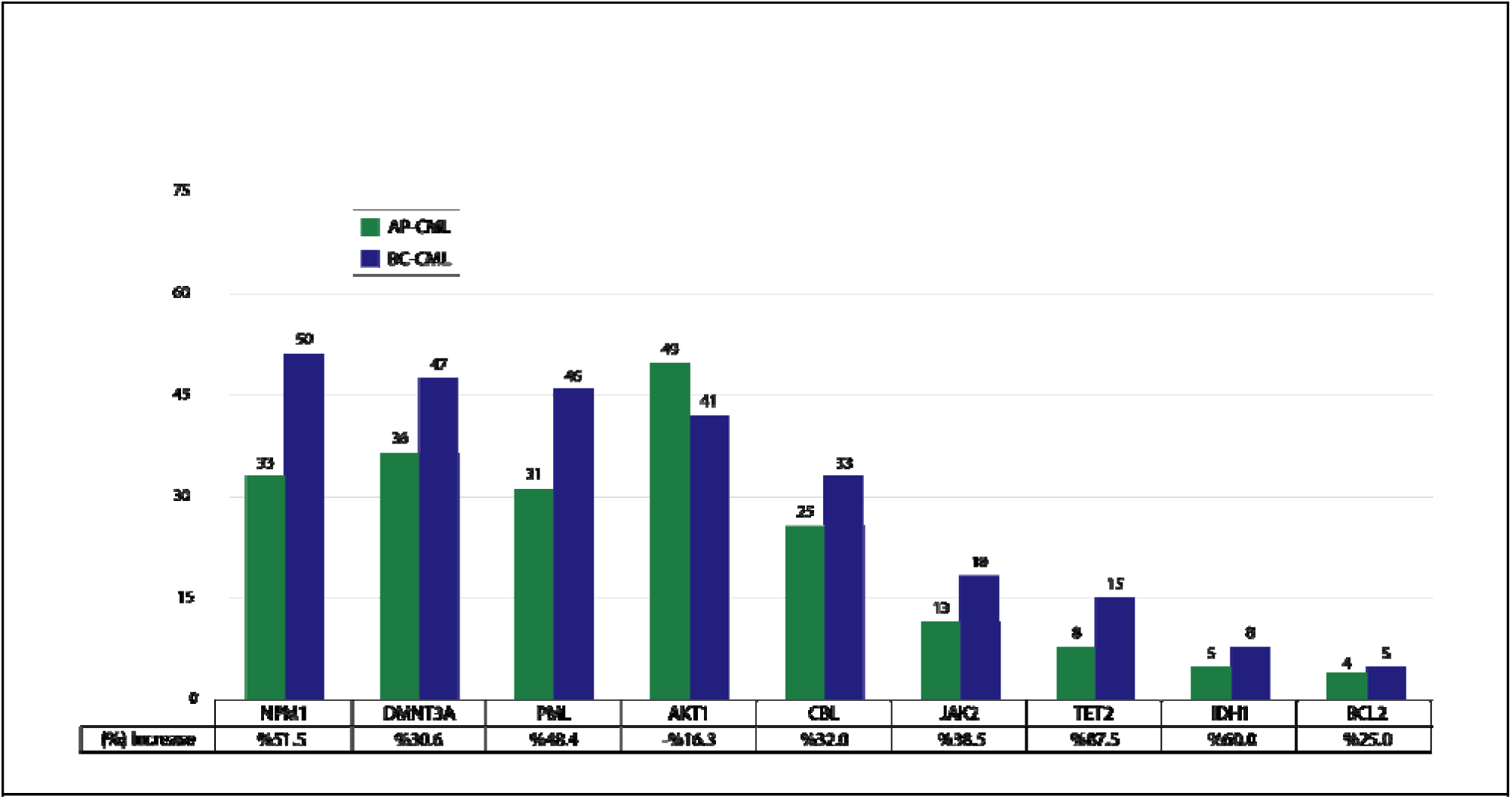
Variants in the druggable genes which have FDA approved drugs to treat AML or ALL and their percentage increase from AP-CML to BC-CML patients in our study subjects.

**Table 2:** Druggable genes with FDA approved drugs to treat AML or ALL.

| Gene | Drug | FDA approved | Clinical trial | Experimental studies | Interaction |
| --- | --- | --- | --- | --- | --- |
| **AKT1** | ARSENIC TRIOXIDE | APL | AKT1-APL |  | direct target |
| **BCL2** | VENETOCLAX | AML (combined with azacitidine, decitabine, or low-dose cytarabine) & CLL with 17p deletion | CML |  | direct target |
|  | DOXORUBICIN | AML & ALL | BCL2-AML |  | biomarker |
|  | MITOXANTRONE | AML | BCL2-AML |  | biomarker |
|  | TRETINOIN (ATRA) | APL | BCL2-AML |  | biomarker |
|  | VINCRISTINE | ALL & lymphoid BC-CML | BCL2-ALL |  | biomarker |
| **CBL** | QUIZARTINIB | FLT- AML | CBL-AML |  | biomarker |
| **DNMT3A** | DECITABINE | CMML (combined with cedazuridine) & AML | DNMT3A-AML |  | direct target |
|  | AZACITIDINE | AML | DNMT3A-AML |  | biomarker |
| **IDH1** | IVOSIDENIB | IDH1-AML | - |  | direct target |
|  | AZACITIDINE | AML | IDH1-ALL (combined with VENETOCLAX ) |  | biomarker |
| **JAK2** | PACRITINIB | Myelofibrosis | - | AML | direct target |
| **NPM1** | VENETOCLAX | AML (combined with azacitidine, decitabine or low dose cytarabine) & CLL with 17p deletion | NPM1-AML |  | biomarker |
| **PML** | ARSENIC TRIOXIDE + TRETINOIN (ATRA) | PML-RARA-APL | - |  | direct target |
| **TET2** | DECITABINE | CMML (combined with cedazuridine) & AML | TET2-CML |  | biomarker |
|  | AZACITIDINE | AML | TET2-CML |  | biomarker |

**Table 3:** FDA approved drugs reported in pandrugs2 to directly target gene mutations found in our cohort and their FDA approved indications.

| Drug | FDA approval | Gene targets (pandrugs) |
| --- | --- | --- |
| VENETOCLAX | AML (22) | BCL2, BCR, EZH2, IDH1 and NPM1 |
| TRETINOIN | APL (23) | BCL2, PDK4, PML and RARA |
| BORTEZOMIB | MM and MCL (24) | BCL2, FGFR3 and GATA2 |
| ZANUBRUTINIB | CLL and SLL (25) | EGFR and JAK2 |
| IVOSIDENIB | IDH1-AML (26) | IDH1 |
| TAZEMETOSTAT | EZH2-Follicular lymphoma (27) | EZH2 |
| RUXOLITINIB | Myelofibrosis (28) | JAK2 and MPL |
| PACRITINIB | Myelofibrosis (29) | JAK2 |
| ARSENIC TRIOXIDE | APL (30) | AKT1 and PML |
| TRETINOIN (ATRA) | APL (23) | BCL2, PDK4, PML and RARA |

**Table 4:**
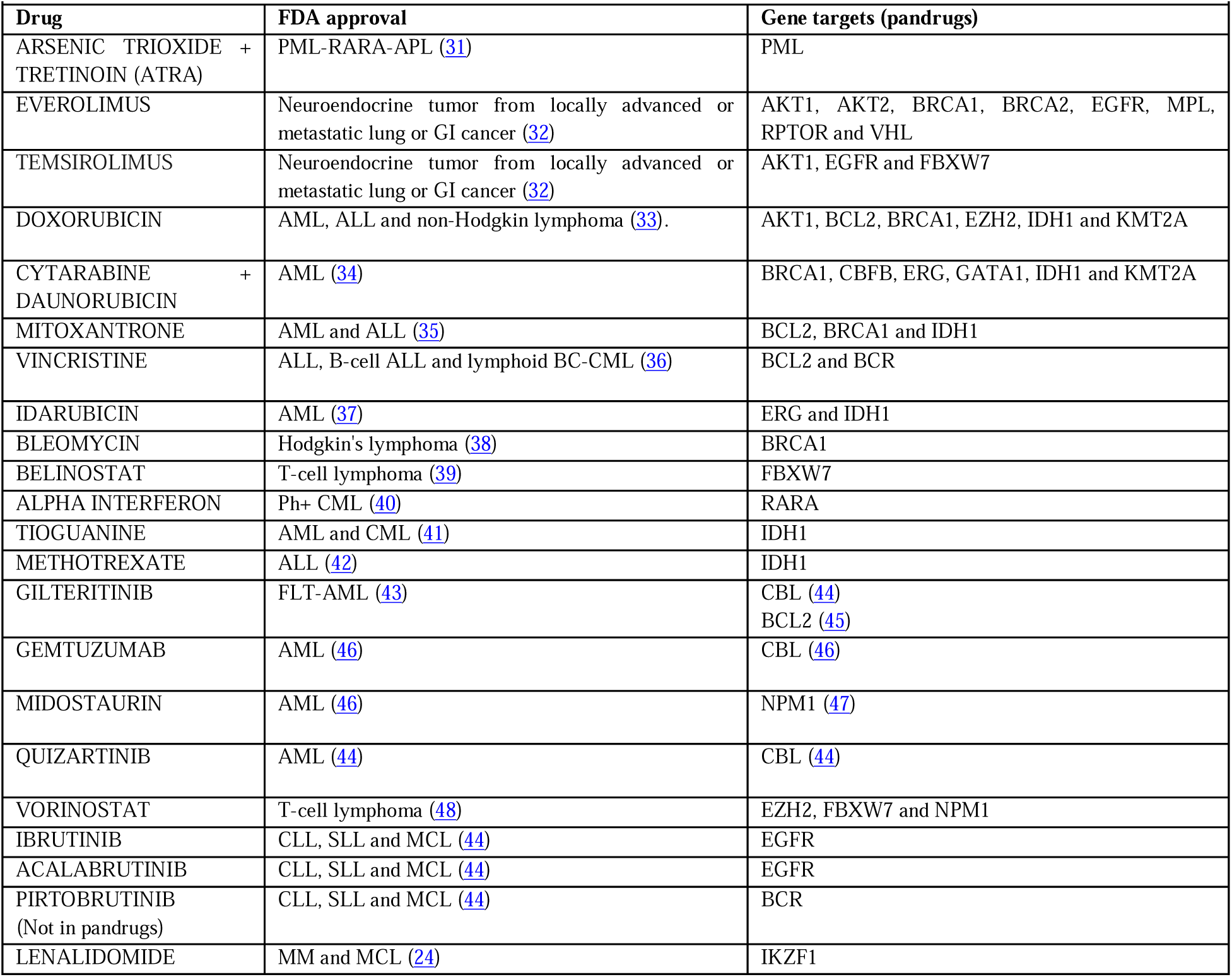
FDA approved drugs reported in pandrugs2 to have a biomarker interaction with gene mutations found in our cohort and their FDA approved indications.

## Repurposing of FDA approved drugs to treat different types of blood cancer using pandrugs2 database and published papers

Overall, significant genetic changes were observed in patients who have progressed to advanced phases of CML. Laboratory findings and clinical characteristics associated with CML progression were anemia, leukocytosis, splenomegaly and hepatomegaly. BC-CML patients had a higher number of variants in their mutated genes, poor treatment response and low overall survival. Genes with the highest variant frequencies in the advanced phases of CML were shared with AML/ALL lineages, except for the ACIN1 gene which was not reported. The druggable genes in AML/ALL lineage genes shared with our samples had low frequencies and variable increase percentages, with TET2 (87.5%) and IDH1 (60.0%) being the highest. Furthermore, IDH1 was the only gene that had an FDA approval drug specifically targeting. Repurposing of these drugs for BC-CML patients should be considered, especially EGFR targeted drug, Zanubrutinib.

Therefore, it is concluded that a large number of AML/ALL-lineage gene mutations are present in BC-CML patients, and FDA-approved and experimental drugs under clinical trials developed against AML- and ALL-specific gene mutations are viable options as potential therapeutics for patient-tailored therapy of BC-CML in TKI- and post-TKI era.

## Discussion

We carried out an extensive investigations on clinical characteristics and AML/ALL lineage gene mutations in BC-CML, at an aim to have a deep insight into biology and genetics of this fatal clinical manifestation in this treatable cancer (CML) (4–8). One of the important findings in the current study was the significant association of BC-CML with hepatomegaly (P=0.0014) while splenomegaly was observed to demonstrate significance in AP-CML (P=0.0160). Overall, mean hemoglobin level, WBC count, platelet count, hepatomegaly, splenomegaly and survival status of Advanced Phase CML patients were significantly different from those of CP-CML patients. However, as these clinical parameters alone cannot help in the early detection of patients at risk of CML progression as well as in finding new drugs to treat difficult-to-treat BC-CML patients, extensive genetic analysis of these patients was vital to find out molecular biomarkers of early progression and to find druggable gene mutations other than BCR-ABL (49). Whole exome sequencing adopted in our study was used to find out such druggable gene mutations and it was one of the best options for this purpose.

Although, six TKIs, imatinib, dasatinib, nilotinib, bosutinib, ponatinib and asciminib are commercially available and approved for treating CML (3) by the Food and Drug Administration (FDA) and European Medicines Agency (EMA) with distinct recommendations based on factors such as the stage of CML at diagnosis, doses, BCR-ABL mutations and reimbursements (3), approximately 20-30% of patients experience resistance to these inhibitors, resulting in treatment failure and disease relapse, which ultimately leads to BC-CML (1). Moreover, despite the provision of all 1^st^, 2^nd^, 3^rd^ and 4^th^ generation TKIs, median overall survival (OS) and failure-free survival (FFS) of BC-CML are 23.8 and 5 months respectively (5, 50). In addition, some TKIs have been shown to cause liver problems such as hepatotoxicity and hepatomegaly, which was also seen in our TKI treated patients (51). This necessitates the need to find additional drug targets and make some other drugs available for treating BC-CML. This could be achieved through drug repurposing from drugs already in clinical practice for AML and ALL treatment, as BC-CML manifests as Myeloid Blast Crisis (M-BC-CML) or Lymphoid Blast Crisis (L-BC-CML) (49). Therefore, we shortlisted genes already reported in AML and ALL (AML and ALL lineage genes) to check their potential as being druggable as many of the gene mutations in AML and ALL are serving as targets for drugs approved by the FDA and EMA, as well as some experimental drugs.

Approximately, 80% of BC patients are M-BC while 20% are L-BC with B cells being more prevalent than T cells (52). Yet, Patients with L-BC have better outcome than those with M-BC. Next generation sequencing (NGS) panel assessment to identify commonly mutated lymphoid or myeloid genes may also be considered but remains an experimental approach in CML. Nevertheless, combined therapies have been adopted for myeloid versus lymphoid BC-CML, based on treatments for AML and ALL respectively. For M-BC as reported by recent small, single arm studies have combined a second generation TKI or ponatinib with decitabine or azacytidine, with CCyR rates of 33-43% and a median OS of 13.8-27.4 months (53). For L-BC, Strati et al evaluated imatinib (400-800 mg daily) or dasatinib (50-140 mg daily) with hyper fractionated cyclophosphamide, vincristine, adriamycin and dexamethasone (hyperCVAD) in 42 patients with L-BC, CCyR and CHR rates were 90% and 58% respectively (54). Also, in a less intensive approach, Rea et al evaluated IM 800 mg together with vincristine and dexamethasone in 13 lymphoid BP patients. Of the 12 evaluable patients, 11 (91.7%) achieved CHR and 4 (33.3%) achieved CcyR (55).

Our WES results indicated that genes with the highest variant frequencies in BC-CML samples were RPTOR (7.3%), followed by BRCA1/BRCA2 (7.0%), BCR (6.0%), STAB1 (4.6%), NF1 (4.4%), ACIN1 (4.4%), EGFR (3.9%), NDRG2 (3.7%), ERG (3.3%) and MYH11 (3.1%) respectively.

However, druggable genes had low variant frequencies in our study subjects. The druggable gene with the highest variant frequency was NPM1 (1.98%), followed by DNMT3A (1.86%), PML (1.82%), AKT1 (1.62%), CBL (1.30%), JAK2 (0.71%), TET2 (0.59V), IDH1 (0.32%) and BCL2 (0.20%) respectively. The AML lineage genes had nine FDA approved drugs while the ALL lineage genes had only one FDA approved drug. This makes sense as both AML and CML are from the myeloid lineage, having more gene mutations in common compared to ALL. Moreover, Ivosidenib was FDA approved to treat AML patients with IDH1 mutations, hence approved as a targeted drug. Although all other FDA approved drugs found in this study are indicated to treat AML/ALL without a specific gene mutation, they are considered targeted therapy as their mechanism of action involve targeting the genes. This indicates that Ivosidenib and the other FDA approved drugs for AML and ALL should also be considered as targeted therapy for CML patients with such gene mutations.

The FDA approved drugs for AML and ALL gene mutations that are found in our BC-CML patients are Arsenic trioxide (ATO) and all-trans retinoic acid (ATRA), Venetoclax (VEN), Ivosidenib (IVO), Azacitidine (AZA), Decitabine (DEC), Doxorubicin (DOX), Mitoxantrone (MIT), Vincristine (VIN), Quizartinib (QUI) and Pacritinib (PAC). ATO and ATRA have received FDA approval for use in the treatment of acute promyelocytic leukemia (APL) (23, 30). A Chinese study has reported that elevated concentrations of ATO effectively maintain ERK phosphorylation, triggering CDKN1A expression and leading to apoptosis. In contrast, treatments with lower doses of ATO cause an increase in AKT1 expression, which inhibits CDKN1A promoter activity and reduces apoptosis. Therefore, ATO demonstrates its therapeutic efficacy predominantly at higher doses (56). Additionally, CML patients exhibiting gain of function mutations in AKT1 may respond favorably to standard doses of ATO, as their AKT1 expression profiles are similar to those observed with low-dose ATO treatments. Meanwhile, ATRA has shown potential in suppressing BCL2 and MCL1, suggesting its utility as a targeted treatment for CML patients carrying BCL2 mutations (57). Furthermore, the FDA has approved a combined regimen of ATRA and ATO specifically for APL patients harboring the PML-RARA fusion. This fusion protein is a direct target of the combined drugs, which have been associated with high rates of complete remission, enhanced overall survival, sustained, profound molecular responses and minimal risk of relapse (31). Consequently, CML patients with PML alterations may benefit significantly from either combination therapy or monotherapy involving ATRA or ATO.

VEN has been FDA-approved as a targeted therapy that explicitly inhibits BCL2 in patients with AML and those with chromosome 17p deletions in chronic lymphocytic leukemia (CLL) (58, 59). Additionally, the combination of dasatinib and VEN has proven its safety and efficiency for managing CP-CML, achieving response rates comparable to those observed with dasatinib treatment alone (60). This supports the potential role of VEN in treating CML patients who exhibit BCL2 variants and may demonstrate resistance to TKIs. AML patients with NPM1 mutations have also shown particular sensitivity to VEN (61). Moreover, VEN has shown remarkable efficacy in conjunction with hypomethylating agents (HMAs) or chemotherapy in treating IDH1-mutated AML and it has effectively addressed relapse in an ALL case involving an IDH1 mutation (62). These findings suggest that VEN could benefit CML patients harboring IDH1, NPM1, or BCL2 variants. Another IDH1-targeted drug that the FDA has approved is IVO, which is indicated for relapsed or refractory AML patients (8). FDA has approved AZA as a maintenance therapy for adult AML patients who cannot undergo further intensive curative treatments and have attained complete remission either with or without incomplete blood count recovery after intensive induction chemotherapy (63). Demonstrating its efficacy, AZA has been used effectively in conjunction with targeted therapies such as VEN, targeting variants in BCL2 and IDH1 (64). Another paper mentioned that old AML patients or those unsuitable for intensive chemotherapy, VEN with HMAs, such as AZA, has not only improved OS but has also established a new standard of care, surpassing outcomes with HMAs alone (65). Furthermore, the FDA has approved the combination of AZA and IVO for treating newly diagnosed IDH1-mutated AML patients, including those who are elderly or have comorbidities (26). In addition, the inclusion of AZA in chemotherapy regimens has extended survival in AML patients specifically harboring a specific DNMT3A variant, R882, thereby suggesting its utility as a potential biomarker for AZA responsiveness in chemotherapy without the need for other targeted therapies (66). AZA’s broad applicability and proven efficacy underscore its potent therapeutic impact in leukemia management and its potential as a targeted treatment for CML patients with mutations in DNMT3A, IDH1, or BCL2, whether as a single agent or in combination with other drugs. DEC is an HMA and DNMT inhibitor that received FDA approval for use in combination with VEN to treat newly diagnosed AML patients who are either elderly or unfit for intensive therapy (61). Furthermore, HMAs have proven effective in patients with TET2 variants (67). DOX, a member of the anthracycline class of chemotherapeutic agents, has been approved by the FDA to treat various cancers, including ALL and AML (68). The drug’s concentration was inversely related to the expression of a BCL2 variant during treatment. Notably, DOX demonstrates a selective capability to induce apoptosis in relapsed AML cells, partly attributed to its targeted inhibition of the BCL2 variant (69). MIT is FDA-approved for treating adult AML (70). Research has shown that MIT triggers programmed cell death in AML cells, accompanied by marked suppression of BCL2 (71). These results indicate that MIT may specifically target mutated BCL2, suggesting its potential utility in treating CML patients with BCL2 variants. VIN is FDA-approved for treating various malignancies, including ALL, B-cell ALL and lymphoid BC-CML (36). Furthermore, an experimental study has demonstrated that VIN effectively reduces BCL2 protein levels in human ALL cells. This effect is enhanced when VIN is used with Huaier aqueous extract (72).

QUI is an FLT3 inhibitor approved by the FDA for FLT3-positive AML patients (44). Loss-of-function (LOF) variants in CBL are associated with a phenotype that exhibits either hyperactivation or increased sensitivity of FLT3. Studies have shown that QUI effectively inhibits cell growth in wild-type FLT3 cells, which is attributed to compromised cCBL E3 ligase activity due to the LOF variant in CBL (73). These findings suggest that CBL mutations could serve as a biomarker for the efficacy of FLT3 inhibitors and offer a promising therapeutic approach for CML patients with CBL variants. PAC is a TKI with equal efficacy against FLT3 and JAK2 and it is FDA-approved for the treatment of myelofibrosis (MF) (29). Research has shown that PAC effectively blocks the phosphorylation of IRAK1 in AML cells leading to reduced viability and survival in AML cell lines carrying various mutations. In addition, IRAK1 is known to be overexpressed in chronic myelomonocytic leukemia (CMML). Treatment with PAC exhibits both standalone activity in primary CMML cells and synergistic effects when used with AZA. Moreover, PAC broadly suppresses kinase signaling pathways in tumor progression including driver mutations in JAK2. Demonstrating clinical tolerability and effectiveness in conditions like chronic myeloproliferative diseases, PAC stands out for its minimal myelosuppressive and immunosuppressive properties compared to other JAK2 inhibitors that also affect JAK1 signaling (74). These attributes underscore PAC’s potential for integration into existing treatment regimens for CML.

### mTOR inhibitors

RPTOR is part of the mTOR pathway which is targeted by mTOR inhibitors (49). FDA-approved mTOR inhibitors are the rapamycin derivatives, Temsirolimus, Everolimus and Ridaforolimus, but they are not indicated for blood cancer (32). It has been reported that Temsirolimus and Everolimus inhibited mTORC2 in AML cells by activating AKT signaling pathway. They also reported that other clinical studies on hematologic malignancies patients treated with these drugs had the same results (75). These studies suggest that mTOR inhibitors can be used as targeted therapy for leukemia patients with mutations in AKT1, AKT2 and RPTOR. Furthermore, an experimental study has shown that Everolimus was effective in treating TKI resistant CML cells and synergistic effect was observed when it was used with Imatinib (49). Another study demonstrated that ponatinib-resistant CML cells could develop resistance through mechanisms independent of BCR-ABL, primarily involving the activation of the mTOR pathway. mTOR inhibitors were identified as effective agents against these resistant cells, inducing autophagy. In addition, it was found that the inhibition of autophagy, could enhance the cytotoxic effects of mTOR inhibitors in these cells. Specifically, the combination with hydroxychloroquine (HCQ) an autophagy inhibitor, significantly reduced cell viability in vitro and improved survival in vivo compared to treatment with mTOR inhibitor alone (76). The findings suggest the importance of mTOR inhibitors in managing TKI-resistant CML, especially for patients harboring RPTOR variants, which was frequently mutated in our study subjects and encourages further exploration into combination therapies that may enhance the efficacy of existing treatments.

### BTK inhibitors

The B-cell antigen receptor (BCR) signaling is essential for the development of chronic lymphocytic leukemia (CLL). Variants in the BCR gene can affect the signaling pathway and fosters the proliferation and survival of CLL cells. A key component in this pathway is Bruton’s Tyrosine Kinase (BTK), whose inhibition disrupts BCR signaling, reducing CLL cell growth and viability. As such, BTK has become a fundamental target for new therapies in CLL and other B-cell-related disorders. Currently, there are several generations of BTK inhibitors (BTKis) available. Ibrutinib, the first-generation BTKi, has significantly altered the treatment paradigm for CLL, achieving historically high response and survival rates. Second generation BTKis, such as acalabrutinib and zanubrutinib, offer greater BTK specificity, which minimizes side effects. Additionally, the novel BTKi, pirtobrutinib is developed to increase efficacy and address resistance encountered with previous BTKis and it recently has been FDA approved for CLL (77). Using these inhibitors to treat BC-CML with BCR variants should be considered, particularly in combination with TKIs as variants in BCR could be the cause of TKI resistance in these BC-CML patients.

### FLT3 Inhibitors

Gilteritinib and Midostaurin are FLT3 inhibitors which are FDA approved for AML. They are used in various therapeutic regimens showing effectiveness in both newly diagnosed FLT3-mutated AML and in relapsed or refractory scenarios (45). Quizartinib (QUI) is another FLT3 inhibitor approved by the FDA for FLT3-positive AML patients (44). Furthermore, loss-of-function (LOF) variants in CBL are associated with a phenotype that exhibits either hyperactivation or increased sensitivity of FLT3. Studies have shown that QUI effectively inhibits cell growth in wild-type FLT3 cells, which is attributed to compromised cCBL E3 ligase activity due to the LOF variant in CBL (73). A study has demonstrated the role of NPM1 in AML patients as a significant positive prognostic factor in determining the response to Midostaurin (47). These findings suggest that CBL and NPM1 mutations could serve as a biomarker for the efficacy of FLT3 inhibitors and offer a promising therapeutic approach for CML patients with CBL or NPM1 variants.

Furthermore, Xiang, W., et al found that Pyrvinium, an FDA-approved drug for pinworm infections is effective in treating BC-CML patients. Its mechanism of action differs depending on the type of cancer it is used to treat. In the case of CML, Pyrvinium inhibits mitochondrial respiration and combining it with Dasatinib promotes apoptosis and improves the treatment in TKI-resistant CML patients. Nonetheless, it has not been approved for CML patients (78).

## Status of Drug repurposing in evidence-based cancer treatment

Despite considerable progress in hematological oncology, leukemic disorder continues to be a leading cause of death globally posing significant health challenges. Although various treatment schemes exist, these treatments frequently develop drug resistance in malignant cells, reducing anticancer agents’ efficacy. This highlights the urgent need for new therapies to counteract drug resistance. However, it takes 10-15 years and costs 1-2 billion $ to produce a new drug approved for clinical use with a multi-stage process starting with design, until safety and efficacy (79). Moreover, clinical evaluations are needed to determine proper dosing and effectiveness and the transition to clinical use involves meeting numerous regulatory and commercial requirements. In tackling such an obstacle, drug repurposing is implemented, in which repurposing existing medications for alternative therapeutic applications is beyond their original indications. Unlike the traditional de novo drug development process, drug repurposing offers several advantages, including enhanced efficiency, reduced time and financial costs and a diminished risk of failure. Moreover, it capitalizes on existing knowledge of drug mechanisms, thereby bolstering the efficiency of clinical translation (80).

In recent years, significant developments in computational resources facilitated systematic drug repurposing. These resources use various information sources, such as electronic health records, genome-wide association analyses, gene expression profiles, pathway mappings, compound structures, target binding assays and phenotypic profiling data. Additionally, computational repurposing approaches, particularly focusing on machine learning algorithms are being apprised recently. Furthermore, there are databases specifically designed to support in-silico drug repurposing, including Drug Repurposing Hub, repoDB, pandrugs and RepurposeDB (81). Therefore, our findings provide a practical approcah of how integration of genomics / multi-omics technologies and available computational resources can help find novel biomarkers, drug targets and therapeutic options for several clinical manifestations with limited treatment modalities like BC-CML.

## Conclusions

Druggable mutations in genes of the AML/ALL lineage are found in considerable number of BC-CML patients if very sensitive NGS methods are utilized. Our NGA technique with 100X or more coverage can be used to identify druggable gene mutations in AML-/ALL-lineage genes in nearly all BC-CML patients. Drugs corresponding to these specific AML/ALL lineage gene mutations are numerous, and many of them are either in active trials or have already received FDA approval. Therefore, our studies provide practical direction for drug repurposing as well as finding novel treatment options, which can individualize treatment for BC-CML patients. Further large scale and prospective studies involving druggable pan-cancer genes are needed to provide a comprehensive insights of molecular oncogenesis of BC-CML and for finding novel biomarkers, drug targets and therapeutic options for this fatal clinical manifestation in CML.

## Author Contributions

Conceptualization, Z.I., N.A.; methodology, Z.I., B.S.A, F.H.A, A.H.A, S.A. K.A.; software, K.A..; validation, Z.I., K.A.; formal analysis, Z.I., B.S.A, F.H.A, A.H.A, S.A. K.A., A.A.G.,S.A.,S.A.M.,M.A.M., M.A.K, M.F.A.,M.I., A.J., A.H.,D.A., A.A.,G.B.,H.H.A, B.A., S.A.A., M.B., S.A., T.K., F.A.M, Y.S.T, S.S., S.R.M., A.M., S.B., M.A,., A.A., M.S., R.N., I.F.Z.; investigation, Z.I., B.S.A, F.H.A, A.H.A, S.A. K.A…; data curation, Z.I., B.S.A, F.H.A, A.H.A, S.A. K.A..; writing—original draft preparation, Z.I., B.S.A, F.H.A, A.H.A, S.A. K.A..—review and editing, Z.I, N.A., K.A,S.A., A.A, S.A.R, M.R., M.AS.; visualization, Z.I., B.S.A, F.H.A, A.H.A, S.A. K.A…; supervision, Z.I., N.A., S.A., K.A..; project administration, Z.I., N.A., S.A., K.A.; funding acquisition, Z.I., A.M, N.A.,.; All authors have read and agreed to the published version of the manuscript.”

## Funding Information

This work was funded by the National Plan for Science, Technology and Innovation (MAARIFAH), King Abdul-Aziz City for Science and Technology, Kingdom of Saudi Arabia, Grant Number 14-Med-2817-02.

## Institutional Review Board Statement

The study was conducted in accordance with the Declaration of Helsinki. It was approved by Intuitional Review Board (IRB) of King Abdullah International Medical Research Centre (KAIMRC), National Guard Health Affairs, Saudi Arabia through project # RA17/002/A, dated 4^th^ Feb 2019, although no research funding was provided by KAIMRC.

## Informed Consent Statement

Written informed consent has been obtained from the patients to publish this paper.

## Data Availability Statement

Access to data made by next-generation sequencing can be obtained from NCBI, to which it was submitted, at https://www.ncbi.nlm.nih.gov/sra/PRJNA734750 (SRA accession number PRJNA734750; accessed on 21October 2024).

## Data Availability

Access to data made by next-generation sequencing can be obtained from NCBI, to which it was submitted, at https://www.ncbi.nlm.nih.gov/sra/PRJNA734750 (SRA accession number PRJNA734750; accessed on 24 April 2024).

## Acknowledgments

We acknowledge Intuitional Review Board (IRB) of King Abdullah International Medical Research Centre (KAIMRC), National Guard Health Affairs, Saudi Arabia for ethical approval of this project (project # RA17/002/A, dated 4^th^ Feb 2019)., although no research funding was provided. This study was partially supported by the College of Medicine Research Centre, Deanship of Scientific Research, King Saud University, Riyadh, Saudi Arabia. All authors have read and have consented to the acknowledgement.

## Conflicts of Interest

The authors acknowledge no financial or other conflicts of interest.

## Notes

### Competing Interest Statement

The authors have declared no competing interest.

### Author Declarations

The study was conducted in accordance with the Declaration of Helsinki. It was approved by Intuitional Review Board (IRB) of King Abdullah International Medical Research Centre (KAIMRC), National Guard Health Affairs, Saudi Arabia through project # RA17/002/A, dated 4th Feb 2019., although no research funding was provided by KAIMRC.

### Summary of Updates

We have improved the abstract substantially.

